# Risk exposures, risk perceptions, negative attitudes toward general vaccination, and COVID-19 vaccine acceptance among college students in South Carolina

**DOI:** 10.1101/2020.11.26.20239483

**Authors:** Shan Qiao, Cheuk Chi Tam, Xiaoming Li

**Affiliations:** University of South Carolina, Department of Health Promotion Education and Behavior, SC SmartState Center for Healthcare Quality, Columbia, SC, USA

**Keywords:** Vaccine acceptance, risk exposures, risk perceptions, COVID-19, college students

## Abstract

Growing attention has been paid to vaccination in control of the COVID-19 pandemic and young adults is one of the key populations for vaccination. Advanced understanding of young adults’ willingness to take a COVID-19 vaccine and the potential factors influencing their vaccine intention will contribute to the development and implementation of effective strategies to promote COVID-19 vaccine uptake among this group. The current study investigated how risk exposures and risk perceptions of COVID-19 (e.g., perceived susceptibility, severity, and fear of COVID-19) as well as negative attitudes toward general vaccination were related to COVID vaccine acceptance among college students based on online survey data from 1062 college students in South Carolina. Hierarchical linear regression was used to examine the association of these factors with COVID-19 vaccine acceptance controlling for key demographics. Results suggested that perceived severity and fear of COVID-19 were positively associated with vaccine acceptance, while higher level of risk exposures (work/study place exposure) and negative attitude toward general vaccination were associated with low vaccine acceptance. Our findings suggested that we need tailored education messages for college students to emphasize the severity of COVID-19, particularly potential long-term negative consequences on health, address the concerns of side effects of general vaccines by dispelling the misconception, and target the most vulnerable subgroups who reported high level of risk exposures while showed low intention to take the vaccine. Efforts are warranted to increase college students’ perceived susceptibility and severity and promote their self-efficacy in health management and encourage them to take protective behaviors including vaccine uptake.

## Introduction

The coronavirus disease 2019 (COVID-19) pandemic caused by SARS-CoV-2 has spread across the world with millions infected and hundreds of thousands dead. The United States has been leading the world in the number of fatalities due to COVID-19 (250,732, as of November 19, 2020) (Johns Hopkins University and Medicine [JHUM], 2020a). The pandemic has been profoundly and adversely impacting various aspects of our societies from health systems, economic growth, to individual life. In addition to public prevention efforts, developing a safe and efficacious vaccine could be one of most promising strategies to curtail the virus, save lives and end the public health crisis. Researchers around the globe have been working collaboratively for this goal with several vaccine candidates tested in clinical trials showing promising efficacy (Centers of Disease Control and Prevention [CDC], 2020; National Institutes of Health [NIH], 2020). However, the success of COVID-19 vaccination program will largely depend on people’s acceptance of the vaccine. A recent global survey suggested that nearly 30% participants would be hesitate to take a COVID-19 vaccine when it is available (Kwok, Lai, Wei, Wong, & Tang, 2020). Extant literature has explored vaccine acceptance and identified a few demographic and psychosocial correlates such as gender, age, trust in research, knowledge and concerns about the novel vaccine, as well as people’s judgment and perceptions regarding risk of COVID-19 (Grech et al., 2020; Harapan et al., 2020; Palamenghi et al., 2020; Wong et al., 2020).

Risk exposure to the disease is one of essential issues that directly shape people’s assessment of their vulnerability and risk. In the context of COVID-19 pandemic, exposures to crowds and/or people infected by the virus without appropriate preventive measures increase the probabilities of infections. Even being weaponed with personal protective equipment, healthcare providers and other essential workers are considered to have high risk exposures to COVID-19 and given priority in vaccine allocations (National Academies of Sciences and Medicine, 2020; World Health Organization [WHO], 2020). Several studies suggest that being healthcare workers or being involved in the care of COVID-19 patients is positively associated with COVID-19 vaccine acceptance (Detoc et al., 2020; Dror et al., 2020; Harapan et al., 2020; Wang et al., 2020).

Risk perceptions are important precursors to health-related behaviors and other behaviors for dealing with or preventing risks (Paek & Hove, 2017). Risk perceptions refer to individuals’ subjective assessments and appraisals about the probability of experiencing harms or hazards such as injury, illness, and death. Risk perceptions are often composed of two main domains: the cognitive domain which is about how much people understand risks (e.g., perceived susceptibility, perceived severity), and emotional domain which relates to how people feel about risks (e.g., fear, dread). Many health behavior theories such as Health Belief Model (HBM), Protection Motivation Theory (PMT), and Risk Perception Attitude (RPA) models emphasize the rational and cognitive aspects of risk perceptions (Janz & Becker, 1984; Rogers, 1983; Rimal & Real, 2003). Meanwhile, as emotional aspects of risk perception being explored, some hypotheses predict that emotional reactions to risks (e.g., fear about the disease) could be independent of cognitive judgment and even are stronger determinants of individual behaviors (Loewenstein, Weber, Hsee, & Welch, 2001). In the context of COVID-19 vaccine, several studies suggest that perceived susceptibility (likelihood to get infected) and perceived severity are predictors of intention to take a vaccine (Dror et al., 2020; Graffigna, Palamenghi, Boccia, & Barello, 2020). One study suggests that a higher level of fear about COVID-19 is related to higher vaccine acceptance (Detoc et al., 2020).

Beside factors specific to the COVID-19 vaccine, individuals’ attitudes towards general vaccination may also affect their COVID-19 vaccine acceptance. Existing literature has suggested that negative attitudes toward general vaccination remain a barrier to population level inoculation against a number of highly infectious diseases (Brewer, Chapman, Rothman, Leask, & Kempe, 2017). The United States has witnessed a growing anti-vaccine movement and a corresponding reduction in population immunity over the past decade (Piltch-Loeb & DiClemente, 2020). Despite of the extraordinary speed of scientific discovery regarding COVID-19, there are still a lot of uncertainties and concerns on novel vaccines in terms of safety and effectiveness. In such a context, the negative attitude toward vaccination could be exaggerated and impede people’s willingness for COVID-19 vaccine uptake.

College students in Southern states in the United States could be one of target populations for COVID-19 vaccination for several reasons. First, the COVID-19 epidemic is more prevalent and serious in the South. For example, as of October 29, 2020, South Carolina has reported cumulative COVID-19 cases of 173,491, and a testing positivity rate (for least 14 days) of 5.9%, which is beyond the threshold of 5% recommended by WHO for reopening (JHUM, 2020b). Second, most of the college and universities in the South reopened campus after the summer and college students are vulnerable for SARS-CoV-2 infection given most of them are living with roommates in apartments or residence on campus provided by the universities. Third, even there may be some travel restriction policies, colleges students have the needs to travel between their homes and schools, which makes them be more exposed to risk of infection. Fourth, young adults in the US has been identified to have low vaccination coverage. The US national data from 2010 to 2020 have revealed that younger adults (aged 18 to 49) had significantly lower influenza vaccination coverage than other age groups (Centers for Disease Control and Prevention, 2020). To address both the needs and challenges for COVID-19 vaccination among college students, it will be very helpful to understand how risk perceptions of this group may influence their COVID-19 vaccine acceptance. For example, young adults (aged 18 to 30) may develop an optimistic bias believing that they have a lower risk of infection or risks pose a less serious threat to themselves than to other age groups (Pasion, Paiva, Fernandes, & Barbosa, 2020; Wise, Zbozinek, Michelini, Hagan, & Mobbs, 2020). In addition, their decision-making and assessment of risk may be more influenced by their emotional reactions compared to adult counterpart. However, many existing studies on COVID-19 vaccination focus on samples composed of various age groups or healthcare providers, there is a dearth of empirical evidence to explore factors that may affect acceptance of COVID-19 vaccination among college students in the Southern States heavily hit by the COVID-19 pandemic.

Therefore, the current study aims to advance our understanding of key factors that may be associated with vaccine acceptance among college students in South Carolina. Specifically, we will explore 1) whether the risk exposures and risk perception of COVID-19 (perceived susceptibility, perceived severity, fear of COVID-19) are associated with vaccine acceptance; and 2) whether the negative attitudes toward general vaccination is also independently associated with COVID-19 vaccine acceptance.

## Methods

### Participants and procedure

Self-administrated and anonymous online survey was conducted through RedCap (a web-based research platform) (Paris & Hynes, 2019) among college students in South Carolina between September and October 2020. Eligibility criteria for participants included: (1) being 18 years of age or older; and (2) being currently a full-time student enrolled in a university in South Carolina. An email invitation was distributed to students Listservs by various colleges and departments on campus. The invitation included a weblink of this survey and an online consent providing information regarding study purposes, procedure, voluntary nature, and confidentiality. The survey typically took about 20 minutes to complete. The students were encouraged to share the invitation or weblink to other students. All participants were provided with an option (by submitting their email address) to enter a prize drawing to win a $25 Amazon e-gift card. A total of 1370 college students responded to the survey with 308 participants of them completed less than half of the survey, which yields a sample size of 1062 in final data analysis. A total of 974 students entered the prize drawing and 10 e-gift cards were given away through random drawing. The study protocol was approved by the Institute Review Board at University of South Carolina.

### Measures

#### Demographics

Participants were asked for key demographic information including gender, age, race/ethnicity, college year, university affiliation, and annual family income. Because certain categories included very few participants (< 5%), we dichotomized gender (0 = Female, 1 = Male) and race/ethnicity (0 = non-Caucasian, 1 = Caucasian) for data analyses.

### COVID-19 vaccine acceptance

One question was used to assess participants’ likelihood to take a COVID-19 vaccine in the future (i.e., “How likely will you get a COVID-19 vaccine when it is available”). Participants responded this question on a five-point Likert scale (1 = definitely not take it to 5 = Definitely take it). A higher score indicated a greater level of vaccine acceptance.

### COVID-19 risk exposure

A two-item checklist was used to assess whether participants had exposed to situations that may lead to a high risk of COVID-19. One item was related to work/study place exposure (i.e., “studying/working in a crowed space”) and the other was related to personal exposure (i.e., “someone I know has COVID-19”). Participants answered these items using three response options (1 = true, 2 = false, 3 = not sure). In the current study, responses on each item were dichotomized into 0 (false or not sure) and 1 (true) to indicate occurrences of exposure. Sum scores was then generated, with a higher score representing a higher level of COVID-19 risk exposure.

### Risk perceptions of COVID-19

#### Perceived susceptibility of COVID-19

Perceived susceptibility of COVID-19 infection was assessed using a 2-item checklist. The items described scenarios associated with COVID-19 risk due to family medical conditions (“I am at high risk of COVID-19 because of my family medical history”), or personal medical conditions (“I am at high risk of COVID-19 because my own underlying medical conditions”). Participants responded items on a three-option scale (1 = true, 2 = false, 3 = not sure) to indicate if the scenarios applied to them. In the current study, responses were dichotomized into 0 (false or not sure) or 1 (true) to indicate if participants perceived themselves susceptible to COVID-19. Sum scores was generated, with high scores indicating higher perceived susceptibility of COVID-19.

#### Perceived severity of COVID-19

Participants were asked about their agreement of the statement “COVID-19 is just a flu-like disease and generally harmless”. The response options ranged from 1 (strongly disagree) to 5 (strongly agree). The answer was reverse-coded with a higher score indicating a greater perceived severity of COVID-19.

#### Fear of COVID-19

Participants were asked about their agreement of the statement “People should not be scared of COVID-19”. Five response options ranged from 1 (strongly disagree) to 5 (strongly agree). The answer was reverse-coded with a higher score indicating a greater fear of COVID-19.

### Negative attitudes toward general vaccination

Four items from the Vaccine Acceptance Instrument (Sarathchandra, Navin, Largent, & McCright, 2018) was adapted to assess participants’ attitudes toward general vaccination. Items asked about participants’ attitudes toward potential negative consequences of vaccination, such as “Vaccines have side effects” and “Vaccines cause long-term harms”. Respondents rated each item on a seven-option Likert scale ranging from 1 (strongly disagree) to 7 (strongly agree). Sum scores were generated, with a higher score indicating a higher level of negative attitudes toward general vaccination. The Cronbach’s alpha for the four items was .77 in the current study.

### Data analysis

Data were screened for proper coding, univariate outliers (*z* scores), and normality (kurtosis and skewness). Descriptive statistics were reported for demographic variables, COVID-19 vaccine acceptance, COVID-19 risk exposure, and risk perceptions of COVID-19 (i.e., perceived susceptibility of COVID-19, perceived severity of COVID-19, and fear of COVID-19), and negative attitudes toward general vaccination. Bivariate analyses were employed among study variables using Pearson’s correlation test.

Hierarchical linear regression was utilized to examine the association of COVID-19 risk exposure, risk perceptions of COVID-19, and negative attitudes toward general vaccination with COVID-19 vaccine acceptance, controlling for demographic variables. The regression model was run in three steps with COVID-19 vaccine acceptance as the dependence variable: First, demographic variables were entered into the model as independent variables. Second, four COVID-19 specific independent variables (COVID-19 risk exposure, perceived susceptibility of COVID-19, perceived severity of COVID-19, and fear of COVID-19) were entered into the model. Third, negative attitudes toward general vaccination was included in the final model. Standardized regression coefficients for independent variables and coefficient of determination (*r*^2^) of the model were estimated in each step. Multicollinearity was determined using variance inflation (VIF) analysis, with values lower than 10 suggesting no multicollinearity (Hair, Anderson, Tatham, & Black, 1995). All statistical analyses were conducted using SPSS software version 26.

## Results

### Sample characteristics

As shown in Table 1, participants were 23.83 years of age on average (*standard deviation* [*SD*] = 6.66). Majority of participants were female (79.8%), Caucasian (85.9%), had an annual family income of $50,000 to $100,000. More than half of the participants were undergraduates (17.1% Senior, 12.4% Junior, 12.2% Freshmen, and 10.5% Sophomore).

**Table 1.**
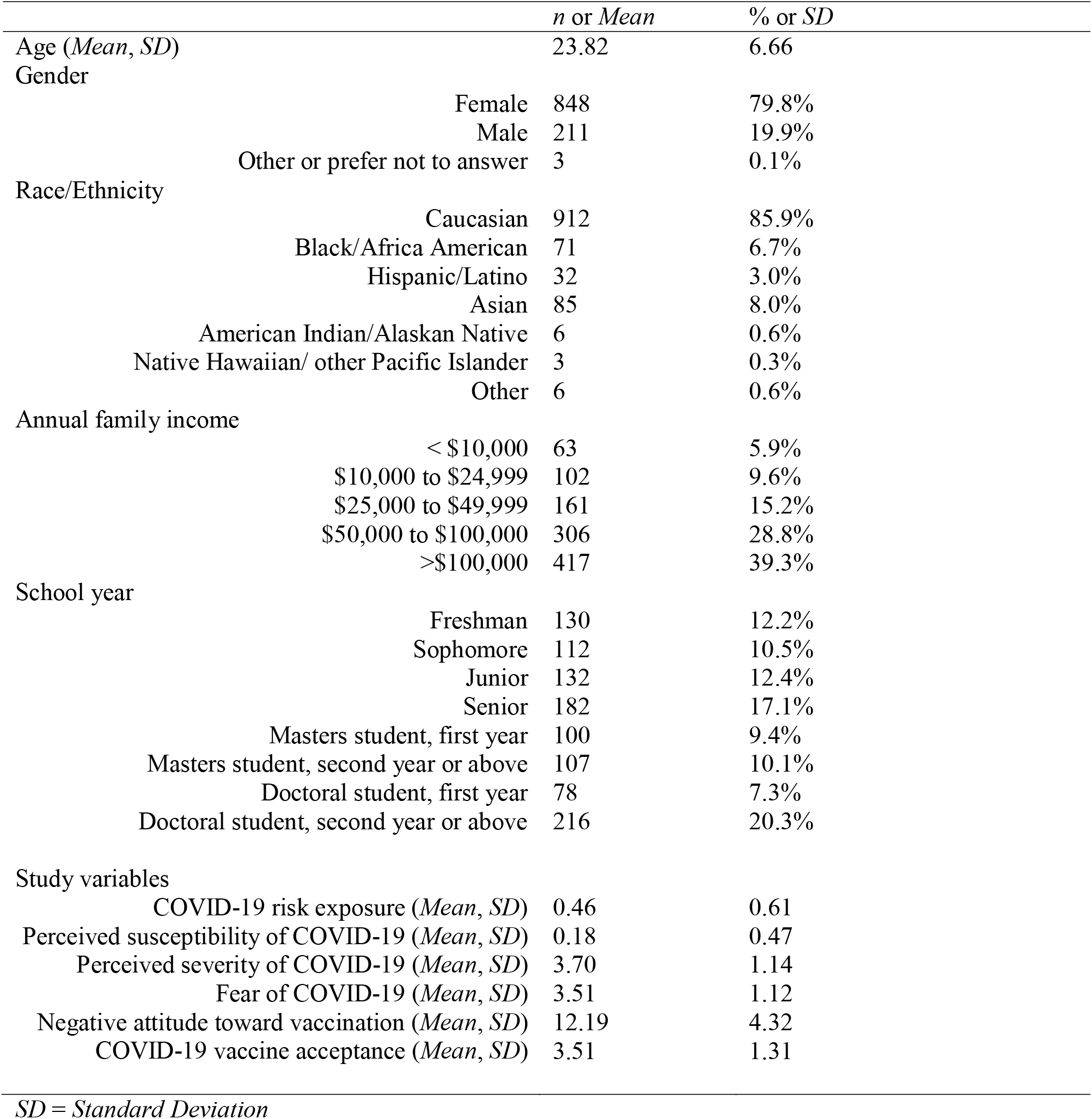
Descriptive statistics of demographic variables and study variables among college students (*N* = 1062)

|  | *n* or *Mean* | % or *SD* |
| --- | --- | --- |
| Age (*Mean, SD*) | 23.82 | 6.66 |
| Gender |  |  |
| Female | 848 | 79.8% |
| Male | 211 | 19.9% |
| Other or prefer not to answer | 3 | 0.1% |
| Race/Ethnicity |  |  |
| Caucasian | 912 | 85.9% |
| Black/Africa American | 71 | 6.7% |
| Hispanic/Latino | 32 | 3.0% |
| Asian | 85 | 8.0% |
| American Indian/Alaskan Native | 6 | 0.6% |
| Native Hawaiian/ other Pacific Islander | 3 | 0.3% |
| Other | 6 | 0.6% |
| Annual family income |  |  |
| < \$10,000 | 63 | 5.9% |
| \$10,000 to \$24,999 | 102 | 9.6% |
| \$25,000 to \$49,999 | 161 | 15.2% |
| \$50,000 to \$100,000 | 306 | 28.8% |
| >\$100,000 | 417 | 39.3% |
| School year |  |  |
| Freshman | 130 | 12.2% |
| Sophomore | 112 | 10.5% |
| Junior | 132 | 12.4% |
| Senior | 182 | 17.1% |
| Masters student, first year | 100 | 9.4% |
| Masters student, second year or above | 107 | 10.1% |
| Doctoral student, first year | 78 | 7.3% |
| Doctoral student, second year or above | 216 | 20.3% |
| Study variables |  |  |
| COVID-19 risk exposure (*Mean, SD*) | 0.46 | 0.61 |
| Perceived susceptibility of COVID-19 (*Mean, SD*) | 0.18 | 0.47 |
| Perceived severity of COVID-19 (*Mean, SD*) | 3.70 | 1.14 |
| Fear of COVID-19 (*Mean, SD*) | 3.51 | 1.12 |
| Negative attitude toward vaccination (*Mean, SD*) | 12.19 | 4.32 |
| COVID-19 vaccine acceptance (*Mean, SD*) | 3.51 | 1.31 |

*SD = Standard Deviation*

### Bivariate analyses

Results of Pearson’s correlations among study variables are presented in Table 2. COVID-19 vaccine acceptance was positively correlated with perceived severity of COVID-19 (*r* = 0.29, *p* < .001), fear of COVID-19 (*r* = 0.31, *p* < .001), and negatively correlated with COVID-19 risk exposure (*r* = -0.07, *p* < .05) and negative attitudes toward general vaccination (*r* = -0.43, *p* < .001). Perceived susceptibility of COVID-19 was positively associated with perceived severity of COVID-19 (*r* = 0.08, *p* < .05) and fear of COVID-19 (*r* = 0.09, *p* < .01). Perceived severity of COVID-19 was positively correlated with fear of COVID-19 (*r* = 0.57, *p* < .001). Lower level of perceived severity (*r* = -0.29, *p* < .001) and fear of COVID-19 (*r* = -0.27, *p* < .001) were correlated with higher negative attitudes toward general vaccination.

**Table 2.** Bivariate analyses among study variables

|  |  | 1 | 2 | 3 | 4 | 5 | 6 |
| --- | --- | --- | --- | --- | --- | --- | --- |
| 1 | COVID-19 vaccine acceptance | 1 |  |  |  |  |  |
| 2 | COVID-19 risk exposure | -0.07* | 1 |  |  |  |  |
| 3 | Perceived susceptibility of COVID-19 | 0.02 | 0.04 | 1 |  |  |  |
| 4 | Perceived severity of COVID-19 | 0.29*** | -0.001 | 0.08* | 1 |  |  |
| 5 | Fear of COVID-19 | 0.31*** | 0.004 | 0.09** | 0.57*** | 1 |  |
| 6 | Negative attitude toward vaccination | -0.43*** | -0.03 | 0.01 | -0.29*** | -0.27*** | 1 |

\* $p < .05$ ; \*\* $p < .01$ ; \*\*\* $p < .001$

**Table 3.** Hierarchical regression on COVID-19 vaccine acceptance among college students

| Predictor Variable | *B* | *SE* | $\beta$ | *t* | *p* | *F* | *R*<sup>2</sup> |
| --- | --- | --- | --- | --- | --- | --- | --- |
| **Step 1 (demographic variables)** |  |  |  |  |  |  |  |
| Gender | 0.24* | 0.11 | 0.07 | 2.21 | 0.027 | 4.06 | 0.02 |
| Age | -0.02** | 0.01 | -0.12 | -3.06 | 0.002 |  |  |
| Race/Ethnicity | 0.09 | 0.12 | 0.02 | 0.71 | 0.479 |  |  |
| Annua family income | 0.03 | 0.04 | 0.03 | 0.81 | 0.416 |  |  |
| School year | 0.06** | 0.02 | 0.11 | 2.84 | 0.005 |  |  |
| **Step 2 (risk exposure and perceptions of COVID-19)** |  |  |  |  |  |  |  |
| Gender | 0.35*** | 0.10 | 0.11 | 3.51 | 0.000 | 19.23 | 0.14 |
| Age | -0.03** | 0.01 | -0.13 | -3.47 | 0.001 |  |  |
| Race/Ethnicity | 0.20 | 0.11 | 0.05 | 1.78 | 0.076 |  |  |
| Annual family income | 0.03 | 0.03 | 0.03 | 0.91 | 0.364 |  |  |
| School year | 0.03 | 0.02 | 0.05 | 1.35 | 0.178 |  |  |
| COVID-19 risk exposure | -0.15* | 0.06 | -0.07 | -2.42 | 0.016 |  |  |
| Perceived susceptibility of COVID-19 | 0.07 | 0.08 | 0.03 | 0.87 | 0.383 |  |  |
| Perceived severity of COVID-19 | 0.19*** | 0.04 | 0.17 | 4.53 | 0.000 |  |  |
| Fear of COVID-19 | 0.28*** | 0.04 | 0.23 | 6.57 | 0.000 |  |  |
| **Step 3 (negative attitude toward vaccination)** |  |  |  |  |  |  |  |
| Gender | 0.23* | 0.10 | 0.07 | 2.47 | 0.014 | 33.36 | 0.24 |
| Age | -0.01* | 0.01 | -0.07 | -2.11 | 0.036 |  |  |
| Race/Ethnicity | -0.02 | 0.11 | 0.00 | -0.14 | 0.889 |  |  |
| Annual family income | 0.01 | 0.03 | 0.01 | 0.19 | 0.848 |  |  |
| School year | -0.02 | 0.02 | -0.03 | -0.78 | 0.436 |  |  |
| COVID-19 risk exposure | -0.17** | 0.06 | -0.08 | -2.78 | 0.006 |  |  |
| Perceived susceptibility of COVID-19 | 0.09 | 0.08 | 0.03 | 1.11 | 0.269 |  |  |
| Perceived severity of COVID-19 | 0.12** | 0.04 | 0.10 | 2.90 | 0.004 |  |  |
| Fear of COVID-19 | 0.20*** | 0.04 | 0.17 | 5.08 | 0.000 |  |  |
| Negative attitude toward vaccination | -0.11*** | 0.01 | -0.35 | -11.71 | 0.000 |  |  |

*SE = Standardized Error*\* $p < .05$ ; \*\* $p < .01$ ; \*\*\* $p < .001$

### Hierarchical linear regression

Table 4 presents results of hierarchical regression analysis. The model 1 explained 2% of variance in COVID-19 vaccine acceptance. Results suggested that college students who were male (*ß* = 0.07, *p* = 0.03), younger (*ß* = -0.12, *p* = 0.002), or at the higher level of school year (*ß* = 0.11, *p* = 0.005) reported higher levels vaccine acceptance.

With risk exposures and risk perceptions of COVID-19 being added, model 2 explained an additional 12% of variance in COVID-19 vaccine acceptance. Results suggested a significantly negative association between COVID-19 risk exposure (*ß* = -0.08, *p* = 0.02) and COVID-19 vaccine acceptance. Perceived severity of COVID-19 (*ß* = 0.17, *p* < 0.001) and fear of COVID-19 (*ß* = 0.23, *p* < 0.001) were positively associated with COVID-19 vaccine acceptance.

The model 3 examined the independent contribution of negative attitudes toward general vaccination beyond the factors in models 1 and 2. The model 3 explained an additional 10% variance of COVID-19 vaccine acceptance compared to the model 2. Results suggested that college students who reported lower levels of negative attitudes toward general vaccination had higher COVID-19 vaccine acceptance (*ß* = -0.35, *p* =< 0.001). All independent variables accounted for 24% of variance COVID-19 vaccine acceptance (*F* = 33.36, *p* < .001). VIF analysis did not show multicollinearity among all independent variables (VIF ranged between 1.03 to 1.73).

## Discussion

The current study is the first one to demonstrate how risk exposures, risk perceptions, and negative attitudes toward general vaccination contribute to COVID-19 vaccine acceptance among college students in a Southern state of United States after controlling for key demographic background. Given college students in this study were generally homogeneous group, their demographic characteristics did not explain much variance (i.e., 2%) of the vaccine acceptance. Risk exposures, risk perceptions, and negative attitudes toward general vaccination together accounted for 22% of variance of the vaccine acceptance suggesting they are all important factors that may influence vaccine acceptance among college students.

Consistent with extant literature on vaccine acceptance, our findings show that higher perceived severity and fear of COVID-19 are associated with higher vaccine acceptance. Although relatively severe consequences of COVID diseases and high mortality rate are reported among the elder population with existing chronic diseases such as severe heart disease, obesity, type 2 diabetes, chronic kidney disease and an immunocompromised condition, there is still so much uncertainty and knowledge gaps about its treatment and care. However, there is some evidence indicating potential prolong illness caused by COVID-19 among young adults and death caused by COVID-19 may occur to a young person too (Tenforde et al., 2020). The perceived COVID-19 severity could increase the willingness to vaccinate against COVID-19 across age groups (Graffigna et al., 2020; Reiter, Pennell, & Katz, 2020). Emotional dimension of risk may affect young people’s decision-making and health-related behaviors. For college students, the fear of the infection may be caused by not only potential clinical outcomes but also social consequences such as social isolation and stigma from their peers.

Existing health behavioral theories posit that higher perceived susceptibility predicts stronger motivations of taking protective actions, such as vaccine uptake. Inconsistent with extant literature on COVID-19 vaccine acceptance, we found that perceived susceptibility was not significantly associated with vaccine acceptance among college students. This may be attributed to the optimistic bias in college students who are not reviewing the virus as a serious threat. Future studies are needed to further explore the different roles of the three risk perception domains in vaccine acceptance and why the impact of perceived susceptibility is not as significant as the other two factors in predicting COVID-19 vaccine acceptance.

The current study confirmed that negative attitudes toward general vaccination such as side effects and potential long-term harm independently contributed to vaccine hesitancy. The concerns on the safety of general vaccines could be intensified when people are facing a new virus. For example, the vaccination rate for the anti-H1N1 vaccine in the 2009 influenza pandemic was below expectations (Lu et al., 2010). The reports of side-effects of some on-going tested vaccine may further impede people’s confidence in vaccine. Surveys suggest that the level of reluctance to vaccinate against COVID-19 is higher than vaccine hesitancy for usual vaccines in several countries (Feleszko, Lewulis, Czarnecki, & Waszkiewicz, 2020). Given the low levels of influenza vaccine acceptance among young adults in the United States (Centers for Disease Control and Prevention, 2020), it is a large challenge to change their attitudes toward vaccination and enhance COVID vaccine uptake.

It is notable that risk exposures were negatively associated with vaccine acceptance, which means, students who reported low level risk exposure showed high vaccine acceptance, while students highly exposed to risks showed low acceptance toward COVID-19 vaccine. There are two possible explanations for this finding. First, the college students who reported low risk exposure were the ones who followed the preventive guideline and more engaged in protecting themselves from the COVID-19 because they avoided working in a crowded space or interacting with a large number of people. It is not surprising that these students demonstrated higher willingness to be vaccinated as an approach to protect themselves against infection. Second, some of the colleges students with a high level of risk exposure might have downplayed the risk of COVID-19 based on their personal experience (as especially as most of them have not been infected) and did not perceive the necessity of a vaccine. Compared with healthcare providers and other essential employees, colleges students may have more controllability over their work environment (e.g., avoid crowds, take on-line classes) and the self-reported low risk exposure may imply their controllability over environment and self-efficacy in health management. Graffigna and colleagues reported that health engagement, individual proactivity in the management of health-related issues, predicated people’s intention to take COVID-19 vaccine across age groups (Graffigna et al., 2020). However, the generally low COVID-19 fatality among young adults may also generate a perception of low risk or invulnerability among college students and lead to a low level of vaccine acceptance. Future studies are needed to further explore the complicated associations between risk exposures, controllability of personal health behavior, and vaccine acceptance.

There are several methodological limitations in the current study. First, the participants were recruited using a convenience sampling approach. Cautions are needed in applying the results in other settings or other populations since correlates of vaccine acceptance could be population and context specific. Second, cross-sectional data would prevent us from drawing any causal inference between risk perception variables and COVID-19 vaccine acceptance. Third, the self-report survey may be subject to response bias (e.g., social desirability). Forth, some measures in the current study were self-developed and have not yet been validated. Future research may benefit from using a random sampling approach, applying a longitudinal design, recruiting college students across different colleges, and validating self-developed measures.

Despite of these limitations, our study explored the impacts of various key perceptions and attitudes on vaccine acceptance among college students confirming the complexity of vaccine acceptance. The investigation of psychosocial reasons of young people that may hamper or promote motivations of getting vaccinated may have some important implications in public health practice since they have been shown as the most hesitant age group for the future COVID vaccine (Barello, Nania, Dellafiore, Graffigna, & Caruso, 2020; Neumann-Böhme et al., 2020). First, we need to tailor the health communication message to young adults emphasizing the severity of COVID-19 disease and its potential long-term negative consequences on health, even for young people. Educational campaigns should be tailored for young adults with comparative optimism and heightened sense of invulnerability. Second, negative attitudes toward general vaccination should be changed by sufficiently addressing the concerns and worries regarding the safety issues. Our findings reveal the importance to create and foster not only positive vaccine-specific attitudes but also general trust and positive attitudes towards vaccination and public health among young generation. Tremendous efforts and effective strategies are warranted to dispel misinformation, misconceptions, and various conspiratorial theories around vaccination. Third, it is urgent for healthcare providers to identify and target the most vulnerable sub-group among college students, those reported high risk exposures and low vaccine acceptance. They may be unable to realize the severity of COVID-19 or lack of controllability/self-efficacy of managing their own health. Interventions are needed to increase their perceived severity and promote their self-efficacy in health management and encourage them to take protective behaviors including vaccine uptake.

## Conclusion

As extant studies suggest, the reluctance toward and refusal of COVID-19 vaccine is a world-wide problem. Different age-group may have different and specific attitudes and concerns regarding the COVID-19 vaccine acceptance. Investigating sociodemographic and psychosocial factors that affect vaccine acceptance is fundamental for an effective immunization plan. The current study provides preliminary findings to understand college students’ acceptance of COVID-19 vaccine and how their risk exposures, risk perceptions and attitudes toward general vaccination contributed to vaccine acceptance. Policy makers and practitioners may target high risk groups by assessing their perceived risks of COVID-19 in cognitive and emotional dimensions, tailor the vaccine communication messages based on college students’ psychological dynamics of decision-making and addressed their concerns, and particularly pay attention to those reported high risks exposures to virus while showed low intention to vaccine uptake.

## Data Availability

Data availability can be contacted through the corresponding author.

## Acknowledgment

We greatly appreciate the advice and comments from Drs. Bankole Olatosi, Sharon Weissman, and Helmut Albrecht on survey development. Research reported in this publication was supported by the National Institutes of Health under Award Number of NIH R01MH0112376-3S1 and R01AI127203-4S1. The content is solely the responsibility of the authors and does not necessarily represent the official views of the National Institutes of Health.

## References

Barello, S., Nania, T., Dellafiore, F., Graffigna, G., & Caruso, R. (2020). ‘Vaccine hesitancy’among university students in Italy during the COVID-19 pandemic. European Journal of Epidemiology, 35(8), 781–783.

Brewer, N. T., Chapman, G. B., Rothman, A. J., Leask, J., & Kempe, A. (2017). Increasing vaccination: putting psychological science into action. Psychological Science in the Public Interest, 18(3), 149–207.

Centers for Disease Control and Prevention. (2020). Different COVID-19 Vaccines. RetreivedNovember 24, 2020 from https://www.cdc.gov/coronavirus/2019-ncov/vaccines/different-vaccines.html

Centers for Disease Control and Prevention. (2020). Flu Vaccination Coverage, United States, 2019–20 Influenza Season. Retrieved November 19, 2020, from https://www.cdc.gov/flu/fluvaxview/coverage-1920estimates.htm

Detoc, M., Bruel, S., Frappe, P., Tardy, B., Botelho-Nevers, E., & Gagneux-Brunon, A. (2020). Intention to participate in a COVID-19 vaccine clinical trial and to get vaccinated against COVID-19 in France during the pandemic. Vaccine, 38(45), 7002–7006.

Dror, A. A., Eisenbach, N., Taiber, S., Morozov, N. G., Mizrachi, M., Zigron, A., … Sela, E. (2020). Vaccine hesitancy: the next challenge in the fight against COVID-19. European Journal of Epidemiology, 35(8), 775–779.

Feleszko, W., Lewulis, P., Czarnecki, A., & Waszkiewicz, P. (2020). Flattening the curve of COVID-19 vaccine rejection—A global overview. Available at SSRN.

Graffigna, G., Palamenghi, L., Boccia, S., & Barello, S. (2020). Relationship between citizens’ health engagement and intention to take the covid-19 vaccine in italy: A mediation analysis. Vaccines, 8(4), 576.

Grech, V., Gauci, C., & Agius, S. (2020). Vaccine hesitancy among Maltese healthcare workers toward influenza and novel COVID-19 vaccination. Early human development, 105213.

Hair, J. F., Anderson, R. E., Tatham, R. L., & Black, W. C. (1995). Multivariate data analysis New York. NY: Macmillan.

Harapan, H., Wagner, A. L., Yufika, A., Winardi, W., Anwar, S., Gan, A. K., … Mudatsir, M. (2020). Acceptance of a COVID-19 vaccine in southeast Asia: A cross-sectional study in Indonesia. Frontiers in Public Health, 8.

Janz, N.K., & Becker, M.H. (1984). The health belief model: A decade later. Health Education Quarterly, 11(1), 1–47.

Johns Hopkins University and Medicine. (2020a). COVID-19 Dashboard by the Center for Systems Science and Engineering (CSSE) at Johns Hopkins University (JHU). Retrieved November 19, 2020, from https://coronavirus.jhu.edu/map.html

Johns Hopkins University and Medicine. (2020b). WHICH U.S. STATES MEET WHO RECOMMENDED TESTING CRITERIA? Retrieved November 19, 2020, from https://coronavirus.jhu.edu/testing/testing-positivity

Kwok, K. O., Lai, F., Wei, W. I., Wong, S. Y. S., & Tang, J. W. T. (2020). Herd immunity– estimating the level required to halt the COVID-19 epidemics in affected countries. Journal of Infection, 80(6), e32–e33.

Loewenstein, G. F., Weber, E. U., Hsee, C. K., & Welch, N. (2001). Risk as feelings. Psychological Bulletin, 127(2), 267.

Lu, P. J., Ding, H., Euler, G. L., Furlow, C., Bryan, L. N., Bardenheier, B., … LeBaron, C. (2010). State-specific influenza A (H1N1) 2009 monovalent vaccination coverage-United States, October 2009-January 2010. Morbidity and Mortality Weekly Report, 59(12), 363–368.

National Academies of Sciences and Medicine, E. (2020). Framework for equitable allocation of COVID-19 vaccine. National Academies Press.

National Institute of Health. (2020). Promising Interim Results from Clinical Trial of NIH-Moderna COVID-19 Vaccine. Retreived on November 26, 2020 from https://www.nih.gov/news-events/news-releases/promising-interim-results-clinical-trial-nih-moderna-covid-19-vaccine

Neumann-Böhme, S., Varghese, N. E., Sabat, I., Barros, P. P., Brouwer, W., van Exel, J., … Stargardt, T. (2020). Once we have it, will we use it? A European survey on willingness to be vaccinated against COVID-19. Springer.

Paek, H.-J., & Hove, T. (2017). Risk perceptions and risk characteristics. In Oxford Research Encyclopedia of Communication.

Palamenghi, L., Barello, S., Boccia, S., & Graffigna, G. (2020). Mistrust in biomedical research and vaccine hesitancy: the forefront challenge in the battle against COVID-19 in Italy. European journal of epidemiology, 35(8), 785–788.

Paris, B. L., & Hynes, D. M. (2019). Diffusion, implementation, and use of Research Electronic Data Capture (REDCap) in the Veterans Health Administration (VA). JAMIA Open, 2(3), 312–316.

Pasion, R., Paiva, T. O., Fernandes, C., & Barbosa, F. (2020). The AGE Effect on Protective Behaviors During the COVID-19 Outbreak: Sociodemographic, Perceptions and Psychological Accounts. Frontiers in Psychology, 11, 2785.

Piltch-Loeb, R., & DiClemente, R. (2020). The Vaccine Uptake Continuum: Applying Social Science Theory to Shift Vaccine Hesitancy. Vaccines, 8(1), 76.

Reiter, P. L., Pennell, M. L., & Katz, M. L. (2020). Acceptability of a COVID-19 vaccine among adults in the United States: How many people would get vaccinated? Vaccine, 38(42), 6500–6507.

Rimal, R.N., & Real, K. (2003). Perceived risk and efficacy beliefs as motivators of change.Human Communication Research, 29(3), 370–399.

Rogers, R.W. (1983). Cognitive and physiological processes in fear appeals and attitude change: A revised theory of protection motivation. In J. Cacioppo & R. Petty (eds.), Social psychophysiology. New York: Guilford.

Sarathchandra, D., Navin, M. C., Largent, M. A., & McCright, A. M. (2018). A survey instrument for measuring vaccine acceptance. Preventive Medicine, 109, 1–7.

Tenforde, M. W., Kim, S. S., Lindsell, C. J., Rose, E. B., Shapiro, N. I., Files, D. C., … Smithline, H. A. (2020). Symptom duration and risk factors for delayed return to usual health among outpatients with COVID-19 in a multistate health care systems network— United States, March–June 2020. Morbidity and Mortality Weekly Report, 69(30), 993.

Wang, K., Wong, E. L. Y., Ho, K. F., Cheung, A. W. L., Chan, E. Y. Y., Yeoh, E. K., & Wong, S. Y. S. (2020). Intention of nurses to accept coronavirus disease 2019 vaccination and change of intention to accept seasonal influenza vaccination during the coronavirus disease 2019 pandemic: A cross-sectional survey. Vaccine, 38(45), 7049–7056.

Wise, T., Zbozinek, T. D., Michelini, G., Hagan, C. C., & Mobbs, D. (2020). Changes in risk perception and self-reported protective behaviour during the first week of the COVID-19 pandemic in the United States. Royal Society Open Science, 7(9), 200742.

Wong, L. P., Alias, H., Wong, P. F., Lee, H. Y., & AbuBakar, S. (2020). The use of the health belief model to assess predictors of intent to receive the COVID-19 vaccine and willingness to pay. Human vaccines & immunotherapeutics, 16(9), 2204–2214.

World Health Organization. (2020). A Global Framework to Ensure Equitable and Fair Allocation of COVID-19 Products and Potential implications for COVID-19 Vaccines. June 18, 2020. Retrieved from https://bit.ly/32rhHPb.

